# Early Detection of Cognitive Decline in Parkinson’s Disease Using Natural Language Processing of Speech: Protocol for a Systematic Review and Meta-Analysis

**DOI:** 10.1101/2025.10.11.25337799

**Authors:** Ravi Shankar, Ziyu Goh, Xu Qian

**Author notes:** **Corresponding Author:** Dr Ravi Shankar; Clinical Research & Innovation Office, Tan Tock Seng Hospital, National Healthcare Group, Singapore, 308433, Email correspondence.

## Abstract

**Background:** Cognitive decline affects up to 80% of Parkinson’s disease (PD) patients, significantly impacting quality of life. Speech changes occur early in PD and may reflect underlying cognitive deterioration before formal diagnosis. Natural language processing (NLP) offers automated, objective methods to analyze speech patterns for cognitive markers, yet evidence remains fragmented across disciplines.

**Objective:** To systematically review and meta-analyze studies using NLP techniques to detect early cognitive decline in PD through speech analysis, evaluating diagnostic accuracy, methodological approaches, and speech-based biomarkers.

**Methods:** Following PRISMA guidelines, we will search PubMed, Embase, Web of Science, IEEE Xplore, PsycINFO, and linguistics databases from inception to December 2025. Eligible studies must apply NLP to speech samples from PD patients for cognitive assessment. Two reviewers will independently screen studies using Covidence. Quality assessment will employ QUADAS-2 for diagnostic studies and PROBAST+AI for prediction models.

**Data Synthesis:** Narrative synthesis will describe NLP methodologies, speech features, and cognitive outcomes. Meta-analysis using bivariate random-effects models will pool diagnostic accuracy measures where appropriate. Subgroup analyses will examine performance by NLP approach, speech task type, and cognitive domain assessed.

**Discussion:** This protocol establishes methodology for the first comprehensive synthesis of NLP-based speech analysis for cognitive detection in PD, addressing critical gaps in existing reviews and providing a framework for evaluating this emerging diagnostic approach.

## Introduction

Parkinson’s disease (PD) represents a complex neurodegenerative disorder affecting approximately 10 million people worldwide, characterized not only by motor symptoms but increasingly recognized for its cognitive manifestations that profoundly impact patient autonomy and caregiver burden [1]. Cognitive impairment in PD follows a heterogeneous trajectory, with mild cognitive impairment (PD-MCI) present in 25-30% of patients at diagnosis and progressing to Parkinson’s disease dementia (PDD) in up to 80% within 20 years, representing one of the most debilitating non-motor features of the disease [2]. The cognitive profile in PD typically involves executive dysfunction, visuospatial deficits, attention impairment, and eventually memory decline, with considerable individual variation in progression patterns and affected domains [3]. Early detection of cognitive decline enables timely interventions, informs treatment decisions regarding medications that may exacerbate cognitive symptoms, facilitates advanced care planning, and provides opportunities for enrollment in neuroprotective trials when interventions may be most effective [4].

Speech production represents a complex cognitive-motor process requiring coordination of multiple neural networks, making it particularly sensitive to neurodegenerative changes in PD [5]. Beyond the well-documented motor speech symptoms of hypophonia and dysarthria, growing evidence suggests that linguistic and cognitive aspects of speech production are affected early in PD, potentially serving as biomarkers for cognitive decline [6]. Speech analysis offers unique advantages for cognitive assessment as it is non-invasive, can be collected remotely via digital devices, allows for frequent monitoring without practice effects, and captures naturalistic cognitive performance in ecologically valid contexts [7]. The intersection of speech and cognition in PD manifests through various measurable parameters including semantic fluency, syntactic complexity, lexical diversity, discourse coherence, and pause patterns, each potentially reflecting different aspects of cognitive function [8].

Natural language processing has emerged as a powerful tool for automated speech analysis, capable of extracting and quantifying linguistic features that may be imperceptible to human observers yet indicative of underlying cognitive changes [9]. Recent advances in NLP, particularly deep learning approaches and transformer-based models, have dramatically improved the ability to analyze complex linguistic patterns, acoustic-prosodic features, and semantic content in speech, with applications ranging from Alzheimer’s disease detection to psychiatric assessment [10, 11]. In PD specifically, preliminary studies have demonstrated that NLP-derived speech features can differentiate between cognitively intact and impaired patients, predict future cognitive decline, and correlate with neuroimaging markers of cognitive dysfunction [12]. However, the rapidly evolving landscape of NLP applications to PD speech analysis has resulted in fragmented evidence across multiple disciplines including neurology, computational linguistics, speech pathology, and biomedical engineering, necessitating systematic synthesis to understand the current state of knowledge and clinical potential.

The justification for this systematic review emerges from critical gaps in the existing literature that limit clinical translation of NLP-based speech analysis for cognitive assessment in PD. While several systematic reviews have examined speech changes in PD, these have primarily focused on motor speech symptoms without systematic attention to cognitive-linguistic features [13], or have examined cognitive assessment broadly without specific focus on NLP methodologies [14]. A recent scoping review by Chen et al. [15] identified speech-based biomarkers for neurodegeneration but included multiple diseases without PD-specific synthesis and excluded studies using advanced NLP techniques published after 2021. Similarly, the systematic review by Sun et al. [16] examined digital biomarkers in PD but combined speech with other modalities such as gait and handwriting, preventing detailed analysis of speech-specific cognitive markers. Furthermore, existing studies have not systematically evaluated the diagnostic accuracy of NLP approaches using meta-analytic methods, limiting evidence-based conclusions about clinical utility [17].

The methodological heterogeneity in this emerging field presents additional challenges not addressed by previous reviews, as studies vary widely in speech elicitation tasks (spontaneous speech, picture description, reading, verbal fluency), NLP approaches (acoustic analysis, lexical features, semantic measures, deep learning), cognitive outcomes (screening tools versus comprehensive neuropsychological assessment), and validation methods [18]. This heterogeneity, while reflecting the interdisciplinary nature of the field, complicates evidence synthesis and clinical interpretation, requiring a systematic framework for categorizing and comparing different approaches. Moreover, the rapid evolution of NLP technology means that reviews conducted even two years ago may not capture current state-of-the-art methods such as large language models and self-supervised learning approaches that show promise for speech-based cognitive assessment [19].

The clinical relevance of establishing an evidence base for NLP-based speech analysis in PD cognitive assessment cannot be overstated, as current diagnostic approaches rely on episodic neuropsychological testing that is resource-intensive, subject to practice effects, and may miss gradual changes between assessments [20]. The COVID-19 pandemic has accelerated interest in remote assessment tools, with speech analysis offering particular promise for telemedicine applications where traditional cognitive testing may be challenging [21]. Furthermore, the potential for continuous, passive monitoring through smart devices could transform cognitive surveillance in PD from reactive diagnosis to proactive risk stratification, enabling earlier intervention when neuroprotective strategies may be most effective [22].

### Objectives

The primary objective of this systematic review and meta-analysis is to comprehensively evaluate the diagnostic accuracy and methodological approaches of natural language processing techniques applied to speech analysis for detecting cognitive decline in Parkinson’s disease patients, providing evidence synthesis to guide clinical implementation and future research directions. Specific objectives include: (1) systematically identifying and categorizing NLP methods used for speech-based cognitive assessment in PD, including acoustic-prosodic analysis, lexical-semantic processing, syntactic complexity measures, and deep learning approaches, with evaluation of their relative performance and methodological requirements; (2) determining pooled estimates of sensitivity, specificity, and area under the curve for NLP-based detection of mild cognitive impairment and dementia in PD through meta-analysis of diagnostically homogeneous studies; (3) examining which speech features and linguistic markers are most strongly associated with cognitive decline across different cognitive domains and stages of impairment; (4) evaluating the influence of speech task type, language, disease severity, and technical factors on diagnostic performance through subgroup analysis and meta-regression; and (5) assessing methodological quality, risk of bias, and reporting completeness to identify best practices and areas requiring standardization for clinical translation.

### Methods Study Design

This systematic review and meta-analysis protocol has been developed following the Preferred Reporting Items for Systematic Review and Meta-Analysis Protocols (PRISMA-P) 2015 statement and will be registered with the International Prospective Register of Systematic Reviews (PROSPERO) prior to commencing searches [23]. The review will adhere to the Meta-analysis of Observational Studies in Epidemiology (MOOSE) guidelines given the anticipated predominance of observational studies in this field [24]. The protocol incorporates recommendations from the Cochrane Handbook for Systematic Reviews of Diagnostic Test Accuracy to ensure appropriate methods for synthesizing diagnostic accuracy studies [25].

### Search Strategy

A comprehensive search strategy will be developed iteratively through collaboration between the review team and an information specialist experienced in biomedical and technical databases. The strategy will employ a combination of controlled vocabulary and free-text terms across three conceptual domains: (1) Parkinson’s disease (“Parkinson*,” “PD,” “parkinsonism,” “paralysis agitans”); (2) cognitive impairment (“cognit*,” “dementia,” “MCI,” “mild cognitive impairment,” “executive function,” “memory,” “neuropsycholog*”); and (3) NLP/speech analysis (“natural language processing,” “NLP,” “computational linguistics,” “speech,” “voice,” “prosody,” “acoustic,” “linguistic,” “machine learning,” “artificial intelligence”).

Electronic databases to be searched include PubMed/MEDLINE, Embase, Web of Science Core Collection, IEEE Xplore, ACM Digital Library, PsycINFO, LLBA (Linguistics and Language Behavior Abstracts), and the ACL Anthology. Grey literature will be identified through ProQuest Dissertations, conference proceedings (INTERSPEECH, ICASSP, MDS Congress), and preprint servers (medRxiv, bioRxiv, arXiv). No date or language restrictions will be applied to the search, though non-English articles will be assessed for translation feasibility. The search will cover publications from database inception through December 2025.

### Eligibility Criteria

Studies will be included based on the PICOS framework: (P) Population - adults with idiopathic Parkinson’s disease diagnosed according to established criteria (UK PDS Brain Bank, MDS criteria), including patients with and without cognitive impairment; studies with mixed populations must report PD-specific results; (I) Index test - any NLP technique applied to speech samples, including acoustic analysis with linguistic processing, lexical-semantic analysis, syntactic processing, discourse analysis, or multimodal approaches combining multiple speech features; (C) Comparator - validated cognitive assessment as reference standard, including neuropsychological testing, MDS criteria for PD-MCI/PDD, or validated screening tools (MoCA, MMSE) with established cutoffs; (O) Outcomes - diagnostic accuracy measures (sensitivity, specificity, AUC) or statistical associations between speech features and cognitive status; (S) Study design -observational studies (cross-sectional, case-control, cohort), excluding case reports and purely qualitative studies.

Exclusion criteria include: studies analyzing only motor speech features without linguistic/cognitive components; written language analysis without spoken speech; studies without clear cognitive outcome measures; conference abstracts without sufficient methodological detail; animal studies; and reviews without original data.

### Study Selection and Data Extraction

Study selection will be managed using Covidence systematic review software. After deduplication, two reviewers will independently screen titles/abstracts and full texts against eligibility criteria, with conflicts resolved through discussion or third reviewer consultation. A standardized data extraction form will capture: study characteristics (design, setting, sample size), population details (age, disease duration, cognitive status, medications), speech task details (type, duration, recording conditions), NLP methodology (features extracted, algorithms, validation), cognitive assessment (tools, diagnostic criteria, timing relative to speech collection), and performance metrics (diagnostic accuracy, correlation coefficients, prediction metrics).

### Quality Assessment

Risk of bias will be assessed using the Quality Assessment of Diagnostic Accuracy Studies-2 (QUADAS-2) tool for diagnostic accuracy studies and the Prediction model Risk Of Bias ASsessment Tool for Artificial Intelligence (PROBAST+AI) for prognostic studies [26, 27]. Additional considerations specific to NLP studies will include: speech sample representativeness, technical validation procedures, handling of confounders (age, education, language proficiency), and reproducibility of methods. Two reviewers will independently conduct assessments with disagreements resolved through consensus.

### Data Synthesis

Narrative synthesis will systematically describe study characteristics, NLP methodologies, and findings organized by speech feature type (acoustic-prosodic, lexical-semantic, syntactic, discourse-level) and cognitive outcome (MCI detection, dementia detection, domain-specific impairment). Where three or more studies report comparable outcomes, meta-analysis will be conducted using hierarchical bivariate random-effects models for diagnostic accuracy studies, jointly modeling sensitivity and specificity to account for threshold effects [28].

Summary receiver operating characteristic (SROC) curves will visualize overall diagnostic performance. Heterogeneity will be assessed using I^2^ statistics and explored through pre-specified subgroup analyses: NLP approach (traditional features vs. deep learning), speech task (spontaneous vs. structured), cognitive domain assessed, and language of assessment. Meta-regression will examine associations between study characteristics and diagnostic performance when sufficient studies are available. Sensitivity analyses will evaluate the impact of study quality, sample size, and cognitive assessment methods on pooled estimates.

Publication bias will be assessed using funnel plots and Deeks’ test when ≥10 studies are included. The Grading of Recommendations Assessment, Development and Evaluation (GRADE) framework will evaluate certainty of evidence considering risk of bias, inconsistency, indirectness, imprecision, and publication bias [29].

## Discussion

This systematic review protocol addresses a critical gap at the intersection of neurology, speech science, and computational linguistics by establishing rigorous methodology for synthesizing evidence on NLP-based speech analysis for cognitive assessment in Parkinson’s disease. The protocol’s comprehensive approach, spanning multiple databases and incorporating both clinical and technical literature, reflects the inherently interdisciplinary nature of this emerging field while ensuring that relevant evidence from diverse research communities is captured and synthesized appropriately. The development of this protocol itself contributes to the field by establishing a framework for evaluating heterogeneous studies that vary in speech tasks, NLP methods, and cognitive outcomes, providing a model for future systematic reviews in related areas of digital biomarker development.

The methodological decisions embedded in this protocol reflect careful consideration of the unique challenges in synthesizing evidence on NLP applications to clinical problems, where technical innovation often outpaces rigorous clinical validation. The choice to include studies from technical venues such as IEEE Xplore and ACL Anthology alongside traditional medical databases recognizes that valuable methodological advances may be published outside conventional clinical journals, yet these studies may lack the clinical rigor expected in medical research [30]. By applying established quality assessment tools (QUADAS-2, PROBAST+AI) while adding NLP-specific considerations, the protocol balances clinical standards with recognition of the technical complexity inherent in computational approaches to speech analysis.

The protocol’s emphasis on diagnostic accuracy as a primary outcome, while also capturing broader associations between speech features and cognition, reflects pragmatic considerations about clinical utility and evidence synthesis. Diagnostic accuracy metrics provide standardized outcomes that enable meta-analysis and direct comparison with conventional cognitive assessments, essential for understanding whether NLP-based speech analysis can complement or potentially replace current diagnostic approaches [31]. However, recognizing that many studies may report continuous associations rather than diagnostic classification, the protocol’s inclusion of correlation and regression analyses ensures comprehensive capture of the relationship between speech and cognition in PD.

Several anticipated challenges in implementing this protocol warrant discussion, as they reflect broader issues in synthesizing evidence on rapidly evolving technological applications in healthcare. The expected heterogeneity in speech tasks—ranging from spontaneous conversation to structured tasks like picture description or verbal fluency— presents challenges for determining which studies are sufficiently similar to pool in meta-analysis. This heterogeneity, while complicating synthesis, also provides opportunity to examine which speech elicitation methods are most sensitive to cognitive changes, informing recommendations for clinical implementation. Similarly, the evolution of NLP methods from traditional linguistic feature extraction to contemporary deep learning approaches raises questions about the comparability of studies using different technical paradigms, necessitating careful subgroup analysis and potentially separate synthesis of traditional versus modern approaches.

The protocol’s inclusive approach to cognitive outcomes, encompassing both formal neuropsychological assessment and screening tools, reflects the reality of cognitive assessment in PD clinical practice and research. While comprehensive neuropsychological testing remains the gold standard for diagnosing PD-MCI and PDD, many studies rely on screening instruments due to practical constraints, and understanding the performance of NLP methods against different reference standards is crucial for interpreting their clinical utility [32]. The planned subgroup analysis by cognitive assessment type will help determine whether NLP performance varies depending on the rigor of the reference standard, informing interpretation of results and recommendations for future research.

The decision to search databases from inception without date restrictions, while potentially increasing the review burden, is justified by the relatively recent emergence of NLP applications to PD speech analysis and the value of understanding how the field has evolved. Early studies using basic acoustic features provide important context for interpreting contemporary deep learning approaches, and examining temporal trends in methodology and performance may reveal whether technological advances translate to improved diagnostic accuracy. The inclusion of non-English studies, contingent on translation resources, recognizes that valuable research may be conducted in languages other than English, particularly given the global distribution of PD and potential language-specific aspects of speech-based cognitive assessment.

Quality assessment represents a particular challenge in this interdisciplinary field, as studies may excel in technical sophistication while lacking clinical rigor, or vice versa. The protocol’s use of established tools (QUADAS-2, PROBAST+AI) ensures systematic evaluation of clinical and epidemiological quality, while additional NLP-specific criteria address technical considerations such as feature selection, model validation, and reproducibility. This dual approach acknowledges that both clinical validity and technical soundness are necessary for successful translation of NLP methods to clinical practice. The decision not to exclude studies based on quality assessment, instead using quality indicators in sensitivity analysis and GRADE evaluation, reflects recognition that this emerging field may include innovative studies with methodological limitations that nonetheless contribute valuable insights.

The protocol’s approach to data synthesis, combining narrative and quantitative methods, provides flexibility to accommodate the expected heterogeneity while generating pooled estimates where appropriate. The narrative synthesis will be particularly valuable for describing the landscape of NLP approaches and identifying methodological trends, while meta-analysis will provide quantitative estimates of diagnostic accuracy essential for clinical decision-making. The use of bivariate models for diagnostic meta-analysis, explicitly modeling the correlation between sensitivity and specificity, represents current best practice for synthesizing diagnostic accuracy studies and will provide more accurate estimates than separate pooling of sensitivity and specificity [33].

The planned subgroup analyses and meta-regression reflect hypotheses about factors that may influence the performance of speech-based cognitive assessment in PD. The comparison of different NLP approaches (traditional features versus deep learning) will address whether increased model complexity translates to improved diagnostic accuracy, informing decisions about the trade-off between interpretability and performance. Analysis by speech task type will determine whether spontaneous speech, which may better reflect real-world communication, offers advantages over structured tasks that provide more standardized assessment. The examination of language effects recognizes that linguistic features may vary across languages, potentially affecting the generalizability of NLP methods developed in English-dominant research settings.

The protocol’s registration with PROSPERO and adherence to reporting guidelines (PRISMA-P) ensures transparency and reduces risk of selective outcome reporting, particularly important in a field where multiple outcomes and analytical approaches are possible. By pre-specifying methods for study selection, quality assessment, and data synthesis, the protocol provides accountability and enables evaluation of whether the completed review adhered to planned methods. This transparency is especially crucial given the potential clinical implications of the review findings, as healthcare providers and policymakers need confidence in the rigor of evidence synthesis to inform decisions about implementing new diagnostic technologies.

The potential limitations of this protocol should be acknowledged, as they may influence the interpretation and applicability of the systematic review findings. The focus on published studies may miss important negative results or ongoing research, potentially biasing estimates of diagnostic accuracy upward. The rapid pace of NLP development means that by the time the review is completed, new methods may have emerged that substantially advance the field. The protocol’s emphasis on diagnostic accuracy may not fully capture other important considerations for clinical implementation such as acceptability to patients, cost-effectiveness, or integration with existing clinical workflows. These limitations highlight the need for regular updating of the review and complementary research addressing implementation considerations beyond diagnostic performance.

The implications of this protocol extend beyond the specific systematic review it describes, contributing to broader efforts to establish evidence-based frameworks for evaluating digital biomarkers in neurodegenerative diseases. As healthcare systems increasingly explore artificial intelligence and digital health technologies, rigorous systematic reviews are essential for distinguishing promising innovations from premature applications. This protocol provides a template for synthesizing evidence on NLP applications in clinical assessment, applicable to other conditions where speech analysis may provide diagnostic insights such as Alzheimer’s disease, depression, or autism spectrum disorder.

## Data Availability

All data produced in the present work are contained in the manuscript

## Notes

### Competing Interest Statement

The authors have declared no competing interest.

### Funding Statement

This study did not receive any funding

## References

1. Global, regional, and national burden of Parkinson’s disease, 1990-2016: a systematic analysis for the Global Burden of Disease Study 2016. Lancet Neurol, 2018. 17(11): p. 939–953.

2. Aarsland, D., et al., Parkinson disease-associated cognitive impairment. Nature Reviews Disease Primers, 2021. 7(1): p. 47.

3. Goldman, J.G. and E. Sieg, Cognitive Impairment and Dementia in Parkinson Disease. Clin Geriatr Med, 2020. 36(2): p. 365–377.

4. Weintraub, D., et al., The neuropsychiatry of Parkinson’s disease: advances and challenges. Lancet Neurol, 2022. 21(1): p. 89–102.

5. Rusz, J., et al., Quantitative acoustic measurements for characterization of speech and voice disorders in early untreated Parkinson’s disease. J Acoust Soc Am, 2011. 129(1): p. 350–67.

6. García, A.M., et al., Cognitive Determinants of Dysarthria in Parkinson’s Disease: An Automated Machine Learning Approach. Mov Disord, 2021. 36(12): p. 2862–2873.

7. Voleti, R., J.M. Liss, and V. Berisha, A Review of Automated Speech and Language Features for Assessment of Cognitive and Thought Disorders. IEEE J Sel Top Signal Process, 2020. 14(2): p. 282–298.

8. Cao, F., et al., Speech and language biomarkers for Parkinson’s disease prediction, early diagnosis and progression. NPJ Parkinsons Dis, 2025. 11(1): p. 57.

9. Fraser, K., et al., Multilingual Prediction of Alzheimer’s Disease Through Domain Adaptation and Concept-Based Language Modelling. 2019. 3659–3670.

10. Shankar, R., A. Bundele, and A. Mukhopadhyay, A Systematic Review of Natural Language Processing Techniques for Early Detection of Cognitive Impairment. Mayo Clinic Proceedings: Digital Health, 2025. 3(2): p. 100205.

11. Shankar, R., A. Bundele, and A. Mukhopadhyay, Natural language processing of electronic health records for early detection of cognitive decline: a systematic review. npj Digital Medicine, 2025. 8(1): p. 133.

12. Eyigoz, E., et al., Linguistic markers predict onset of Alzheimer’s disease. EClinicalMedicine, 2020. 28: p. 100583.

13. Wright, H. and V. Aharonson, Vocal Feature Changes for Monitoring Parkinson’s Disease Progression-A Systematic Review. Brain Sci, 2025. 15(3).

14. Rice, M., S. Warren, and S. Betz, Language symptoms of developmental language disorders: An overview of autism, Down syndrome, fragile X, specific language impairment, and Williams syndrome. Applied Psycholinguistics, 2005. 26.

15. Chen, S., et al., Review of voice biomarkers in the screening of neurodegenerative diseases. Interdisciplinary Nursing Research, 2024. 3.

16. Sun, Y.M., et al., Digital biomarkers for precision diagnosis and monitoring in Parkinson’s disease. NPJ Digit Med, 2024. 7(1): p. 218.

17. Pascuzzo, R., et al., Editorial: A comprehensive look at biomarkers in neurodegenerative diseases: from early diagnosis to treatment response assessment. Front Aging Neurosci, 2025. 17: p. 1642793.

18. Rohl, A., et al., Speech dysfunction, cognition, and Parkinson’s disease. Prog Brain Res, 2022. 269(1): p. 153–173.

19. Kim, N., M. Homer, and H. Jang, Clinical Application of Large Language Models for Intervention Plan Development in Speech-Language Pathology. Am J Speech Lang Pathol, 2025. 34(4): p. 2098–2114.

20. Skorvanek, M., et al., Global scales for cognitive screening in Parkinson’s disease: Critique and recommendations. Mov Disord, 2018. 33(2): p. 208–218.

21. Motolese, F., et al., Parkinson’s Disease Remote Patient Monitoring During the COVID-19 Lockdown. Front Neurol, 2020. 11: p. 567413.

22. Arora, S., et al., Smartphone Speech Testing for Symptom Assessment in Rapid Eye Movement Sleep Behavior Disorder and Parkinson’s Disease. IEEE Access, 2021. PP: p. 1–1.

23. Shamseer, L., et al., Preferred reporting items for systematic review and meta-analysis protocols (PRISMA-P) 2015: elaboration and explanation. Bmj, 2015. 350: p. g7647.

24. Stroup, D.F., et al., Meta-analysis of observational studies in epidemiology: a proposal for reporting. Meta-analysis Of Observational Studies in Epidemiology (MOOSE) group. Jama, 2000. 283(15): p. 2008–12.

25. Bossuyt, P.M., et al., Evaluating medical tests: introducing the Cochrane Handbook for Systematic Reviews of Diagnostic Test Accuracy. Cochrane Database Syst Rev, 2023. 7(7): p. Ed000163.

26. Whiting, P.F., et al., QUADAS-2: a revised tool for the quality assessment of diagnostic accuracy studies. Ann Intern Med, 2011. 155(8): p. 529–36.

27. Moons, K.G.M., et al., PROBAST+AI: an updated quality, risk of bias, and applicability assessment tool for prediction models using regression or artificial intelligence methods. BMJ, 2025. 388: p. e082505.

28. Reitsma, J.B., et al., Bivariate analysis of sensitivity and specificity produces informative summary measures in diagnostic reviews. J Clin Epidemiol, 2005. 58(10): p. 982–90.

29. Schünemann, H.J., et al., Grading quality of evidence and strength of recommendations for diagnostic tests and strategies. Bmj, 2008. 336(7653): p. 1106–10.

30. Beam, A.L. and I.S. Kohane, Big Data and Machine Learning in Health Care. Jama, 2018. 319(13): p. 1317–1318.

31. Bossuyt, P.M., et al., STARD 2015: an updated list of essential items for reporting diagnostic accuracy studies. BMJ : British Medical Journal, 2015. 351: p. h5527.

32. Litvan, I., et al., Diagnostic criteria for mild cognitive impairment in Parkinson’s disease: Movement Disorder Society Task Force guidelines. Mov Disord, 2012. 27(3): p. 349–56.

33. Takwoingi, Y., R.D. Riley, and J.J. Deeks, Meta-analysis of diagnostic accuracy studies in mental health. Evid Based Ment Health, 2015. 18(4): p. 103–9.

